# Real-world effectiveness of sotrovimab for the treatment of SARS-CoV-2 infection during Omicron BA.2 subvariant predominance: a systematic literature review

**DOI:** 10.1101/2023.03.09.23287034

**Authors:** Myriam Drysdale, Daniel C. Gibbons, Moushmi Singh, Catherine Rolland, Louis Lavoie, Andrew Skingsley, Emily J. Lloyd

**Affiliations:** Value Evidence and Outcomes, GSK, Middlesex, UK; Evidence Synthesis, Modelling and Communications, PPD Evidera, London, UK; Evidence Synthesis, Modelling and Communications, PPD Evidera, Montreal, Canada; Clinical Research and Development, GSK, Middlesex, UK

**Keywords:** COVID-19, Omicron BA.2, Monoclonal antibody, Sotrovimab Hospitalizations, Mortality

## Abstract

**Purpose:** Emerging severe acute respiratory syndrome coronavirus 2 (SARS-CoV-2) variants have impacted the in vitro activity of sotrovimab 500 mg, with reduced fold change in EC_50_ for the Omicron BA.2 sublineage and onward. The correlation between this reduction and clinical efficacy outcomes is unknown. In the absence of clinical trials assessing the efficacy of sotrovimab against emerging variants, real-world evidence becomes a critical source of information. A systematic literature review (SLR) of published observational studies was undertaken to evaluate the effectiveness of sotrovimab on severe clinical outcomes during the Omicron BA.2 subvariant predominance period.

**Methods:** Searches of indexed electronic databases for peer-reviewed journals, preprint articles, and conference abstracts published between January 1, 2022 and November 3, 2022 were undertaken using a combination of search terms for COVID-19, sotrovimab, and observational study design. Study quality was assessed using the Newcastle Ottawa Scale (NOS).

**Results:** From the 343 unique titles and abstracts identified, five studies were eligible for inclusion in the SLR. Included studies displayed heterogeneity in study design and population. The OpenSAFELY study, which received a high NOS score and had a sufficient sample of patients treated with sotrovimab during BA.2 predominance, demonstrated clinical effectiveness during both BA.1 (adjusted hazard ratio (HR) 0.54, 95% confidence interval (CI) 0.33–0.88; *p* = 0.014) and BA.2 (adjusted HR 0.44, 95% CI 0.27–0.71; *p* = 0.001) periods vs molnupiravir. Furthermore, a US-based study that also received a high NOS score reported that sotrovimab was associated with a lower risk of 30-day all-cause hospitalization or mortality compared with no monoclonal antibody treatment during the BA.2 subvariant surge in March (adjusted relative risk (RR) 0.41, 95% CI 0.27–0.62) and April 2022 (adjusted RR 0.54, 95% CI 0.08–3.54). Although only a limited number of studies evaluated sotrovimab during both the BA.1 and BA.2 periods, these demonstrated that clinical outcomes in patients with COVID-19 treated with sotrovimab were consistently low across both periods. One large study directly compared data from the two periods and found no evidence of a difference in the clinical outcomes of sotrovimab-treated patients with sequencing-confirmed BA.1 and BA.2 (HR 1.17, 95% CI 0.74–1.86).

**Conclusion:** The observational data presented in this SLR provide evidence that the effectiveness of sotrovimab (IV 500 mg) is maintained against Omicron BA.2 in both ecological and sequencing-confirmed studies, either through the demonstration of low and comparable rates of severe clinical outcomes between the Omicron BA.1 and BA.2 periods, or by comparison against an active comparator or no treatment within the Omicron BA.2 period.

**Key points:** *Why carry out this study?:*

- Emerging SARS-CoV-2 variants have impacted the in vitro activity of sotrovimab 500 mg, with reduced fold change in EC_50_ relative to wild-type for the Omicron BA.2 sublineage and onward; the clinical relevance of this difference on outcomes for BA.2 (and other variants) is unknown.
- Given the complexity of generating formal clinical trial data in the context of the constantly evolving SARS-CoV-2 landscape, real-world evidence is a key source of information with which to assess the effectiveness of treatments such as sotrovimab on newly predominant or emerging variants.
- We conducted a systematic literature review to evaluate the effectiveness of sotrovimab for the early treatment of COVID-19 on clinical outcomes during the period predominated by the Omicron BA.2 subvariant.

*What was learned from the study?:*

- Sotrovimab treatment was associated with low proportions of severe clinical outcomes (such as all-cause or COVID-19-related hospitalization or mortality) in patients infected during periods of Omicron BA.2 predominance, despite reduction in the in vitro neutralization activity of sotrovimab.
- These data support continued clinical effectiveness of sotrovimab during Omicron BA.2 predominance.

## Introduction

Coronavirus disease 2019 (COVID-19) is caused by infection with severe acute respiratory syndrome coronavirus 2 (SARS-CoV-2). Following its initial emergence in December 2019 and the subsequent declaration of a pandemic by the World Health Organization (WHO) in March 2020 [1], the virus has continued to evolve and continues to place pressure on healthcare systems globally. Some individuals, such as older patients, immunocompromised patients, or those with advanced renal or liver disease, diabetes, cancer, chronic obstructive pulmonary disease, or cardiovascular disease, are at a higher risk of developing severe COVID-19 [2–4].

Clinical outcomes of COVID-19 are influenced by country-level factors such as healthcare system capacity and policies for disease prevention and management, as well as individual-level factors such as age, pre-existing illnesses, and immune status [2, 3, 5, 6]. Moreover, new SARS-CoV-2 variants continue to emerge globally, affecting viral transmissibility, pathogenicity, and antigenic capacity, thus potentially impacting the spectrum and severity of clinical outcomes, immune evasion, and treatment effectiveness in infected individuals [7].

Sotrovimab is a dual-action recombinant human IgG1κ monoclonal antibody (mAb) derived from the parental mAb S309, a potent neutralizing mAb directed against the spike protein of SARS-CoV-2 [8–11]. In a randomized clinical trial (COMET-ICE, NCT04545060) conducted during the period of the pandemic predominated by the original “wild-type” variant, a single intravenous (IV) infusion of sotrovimab (500 mg) was found to significantly reduce the risk of all-cause hospitalization (of >24-hour duration) or death by 79% compared with placebo in high-risk patients with COVID-19 [12, 13]. Consequently, sotrovimab (IV 500 mg) was first granted Emergency Use Authorization (EUA) by the United States (US) Food and Drug Administration for the treatment of mild-to-moderate COVID-19 in adults and pediatric patients (≥12 years of age and ≥40 kg) who tested positive for SARS-CoV-2 and were at a high risk of progression to severe COVID-19, including hospitalization or death [14]. Sotrovimab was then authorized by several regulatory agencies across the world, including the European Medicines Agency [15].

Since the COMET-ICE trial was undertaken, the original “wild-type” virus has evolved, leading to the emergence and establishment of new variants, with the Alpha variant being the first recognized by the WHO as a variant of concern at the end of 2020 [16]. A number of other recognized variants subsequently emerged, including the Omicron BA.2 subvariant that became predominant globally in March 2022 [7, 17]. In vitro neutralization assays demonstrated that sotrovimab retained its neutralization capacity against Omicron BA.1 (3.8-fold EC_50_ reduction relative to wild-type SARS-CoV-2), but showed reduced neutralization against Omicron BA.2, BA.4, BA.5, and BA.2.12.1, with 16-, 21.3-, 22.6-, and 16.6-fold reductions, respectively [18]. In the absence of clinical trials to assess the efficacy of sotrovimab against these emerging variants, the clinical relevance of this reduced neutralization observed in vitro was unknown. In lieu of evidence supporting the efficacy of sotrovimab against BA.2, sotrovimab was de-authorized in the US on a state-by-state basis from the end of March 2022, with a national deauthorization occurring on April 5, 2022 [19].

Considering the rapid rate of SARS-CoV-2 mutation and the ever-evolving variant landscape, the growing body of published real-world evidence is a key source of information with which to assess the effectiveness of sotrovimab on newer variants outside of clinical trials. A published systematic literature review (SLR) and meta-analysis of 17 studies including 27,429 patients concluded that sotrovimab is an effective and well-tolerated therapy that can reduce mortality and hospitalization rates in patients infected with both the Delta (odds ratio [OR] 0.07; 95% CI 0.01– 0.51) and Omicron BA.1 (OR 0.27; 95% CI 0.14–0.51) circulating variants [20].

Despite de-authorization in the US, sotrovimab remained authorized in other countries [15], and use continued for early treatment of COVID-19 in high-risk populations during BA.2 predominance. To address some of the questions regarding the use of sotrovimab against emerging variants, this SLR was undertaken to evaluate the totality of evidence on clinical effectiveness of sotrovimab (IV 500 mg) during the Omicron BA.2 predominance period and onwards.

## Methods

This SLR included observational studies investigating clinical outcomes and viral load in patients treated with sotrovimab published in peer-reviewed journal articles, preprint articles, and conference abstracts between January 1, 2022 and November 3, 2022. Only clinical outcomes were reported for the purpose of this publication. The SLR was conducted in accordance with Preferred Reporting Items for Systematic Reviews and Meta-Analyses (PRISMA) guidelines (PROSPERO registration number: CRD42022376733) [21].

The publication period covered by the systematic review was selected to identify data on Omicron BA.2 and subsequent subvariants. Where available, data on other circulating variants were also extracted for potential comparison between periods of variant predominance.

### Data sources and search strategy

Searches were conducted on November 3, 2022 in the following indexed electronic databases: MEDLINE (via OVID), Embase (via OVID), LitCovid (via MEDLINE), Cochrane COVID-19 Study Register, and EconLit. Additional searches for relevant preprints were conducted in ArRvix, BioRvix (via Embase), ChemRvix, MedRvix (via Embase), Preprints.org, ResearchSquare, and SSRN. The following conferences were also searched for relevant abstracts indexed from January 2022: (1) Infectious Diseases Week, (2) International Conference on Emerging Infectious Diseases, (3) European Respiratory Society, and (4) European Congress of Clinical Microbiology and Infectious Diseases. These conferences were selected as they were likely to include a wide range of newly available research in the field of COVID-19 therapeutics and management.

Search strategies, starting from January 1, 2022 for each database, included a combination of free-text search terms for COVID-19, sotrovimab, and observational study design (Supplementary Table 1). There was no limit on geographical location, but only English language publications were considered.

### Study selection

Studies were screened and selected for inclusion in the SLR against predetermined population, interventions and comparators, outcomes, and study design criteria [22]. Only studies matching any inclusion criteria and none of the exclusion criteria listed in Table 1 were eligible for inclusion. As the focus of this SLR was outcomes captured during Omicron BA.2 predominance, only papers reporting this are included here.

**Table 1.** Inclusion and exclusion criteria

| Domain | Criteria | Exclusion reason | Exclusion description |
| --- | --- | --- | --- |
| Populations | Patients aged $\geq 12$ years who fulfill the following criteria:<br>Identified as having confirmed COVID-19<br>Have received sotrovimab for treatment of SARS-CoV-2 infection as per standard of care<br>Presented with the BA.2 subvariant or had COVID-19 during BA.2 subvariant predominant period<br>Subgroups of interest:<br>Subgroup within high-risk group (i.e. transplant patients, renal patients) | Population not of interest | Patients aged $< 12$ years |
| Interventions/Comparators | All studies on sotrovimab-treated patients ($n \geq 20$) | No treatment of interest | Did not receive sotrovimab<br>Received sotrovimab as a prophylactic treatment, or for primary treatment of severe COVID-19<br>Less than 20 patients treated with sotrovimab |
| Outcomes | Following clinical outcomes within 30 days of sotrovimab:<br>Hospitalization<br>Intensive care admission<br>Emergency department visits<br>Respiratory support (e.g. use of supplemental oxygen)<br>COVID-19 progression (e.g. composite endpoint such as ICU/respiratory support/mortality)<br>Mortality<br>Absolute (change from baseline) and relative change (compared to BA.1 period, active and untreated comparators) in viral load during the acute phase post-sotrovimab | Outcomes not of interest | Relevant outcomes are not reported |
|  | • Proportion of patients with undetectable viral load post-sotrovimab treatment |  |  |
| Study design | Any of the following study designs:<br>• Observational studies (including sotrovimab-treated single-arm studies and comparative effectiveness studies)<br>• SLRs with or without meta-analysis (for citation chasing of observational studies only) | • Publication type not of interest<br>• Study design not of interest | • Case Report, Editorial, Opinion Piece, Letter to the Editor, Clinical Trial, Narrative Review, Guidelines<br>• Pre-clinical studies (animal, in vitro, ex vivo, pharmacokinetics)<br>• Clinical trials |

*COVID-19* coronavirus disease 2019, *ICU* intensive care unit, *SARS-CoV-2* severe acute respiratory syndrome coronavirus 2, *SLR* systematic literature review

Two independent reviewers evaluated each title and abstract against the defined selection criteria to determine suitability for the SLR, and a third reviewer resolved disagreements. The same process was applied for the review of the full-text articles.

### Data extraction and quality assessment

Extraction of data from the included studies was performed by a single extractor using a data extraction file designed in Microsoft Excel. An independent researcher reviewed all extracted fields, and discrepancies were resolved by a third reviewer.

Extracted information included the study title and reference, study details and design, country, data source, study population, number of patients, data collection period and associated circulating SARS-CoV-2 variants, follow-up duration, sponsor, key baseline characteristics, and clinical outcomes. Clinical outcomes included hospitalization, intensive care admission, emergency department visits, respiratory support (e.g. use of supplemental oxygen), COVID-19 progression (e.g. composite endpoint such as intensive care unit [ICU]/respiratory support/mortality), mortality, absolute (from baseline) and relative (from Omicron BA.1 period, active or untreated comparators) change in viral load during the acute phase post-sotrovimab treatment, and proportion of patients with undetectable viral load post-sotrovimab treatment.

The Newcastle Ottawa Scale (NOS) was used to assess the quality of each study by considering characteristics that could introduce bias [23, 24]. Studies were judged on three broad domains of their design: (1) selection of study groups, (2) comparability of groups, and (3) ascertainment of either the exposure or outcome of interest for case-control or cohort studies, respectively. The maximum attainable score in a NOS quality assessment is 9 (accumulated across all domains), with greater scores representing a lower risk of bias.

## Results

### Study selection

Electronic database searches initially yielded a total of 257 papers. An additional 263 studies were obtained from searching conference abstracts, preprints, and citation chasing from relevant SLRs (Fig. 1). After the removal of duplicates, 343 unique titles and abstracts were screened, of which 89 were considered admissible for full-text review. Of these, five observational studies containing clinical or viral load outcome data for sotrovimab from the BA.2 predominance period were considered eligible for inclusion in the SLR [25–29]. We did not identify any studies describing clinical outcomes post-BA.2. Reasons for exclusion during the full-text review are detailed in Fig. 1.

**Fig. 1.**
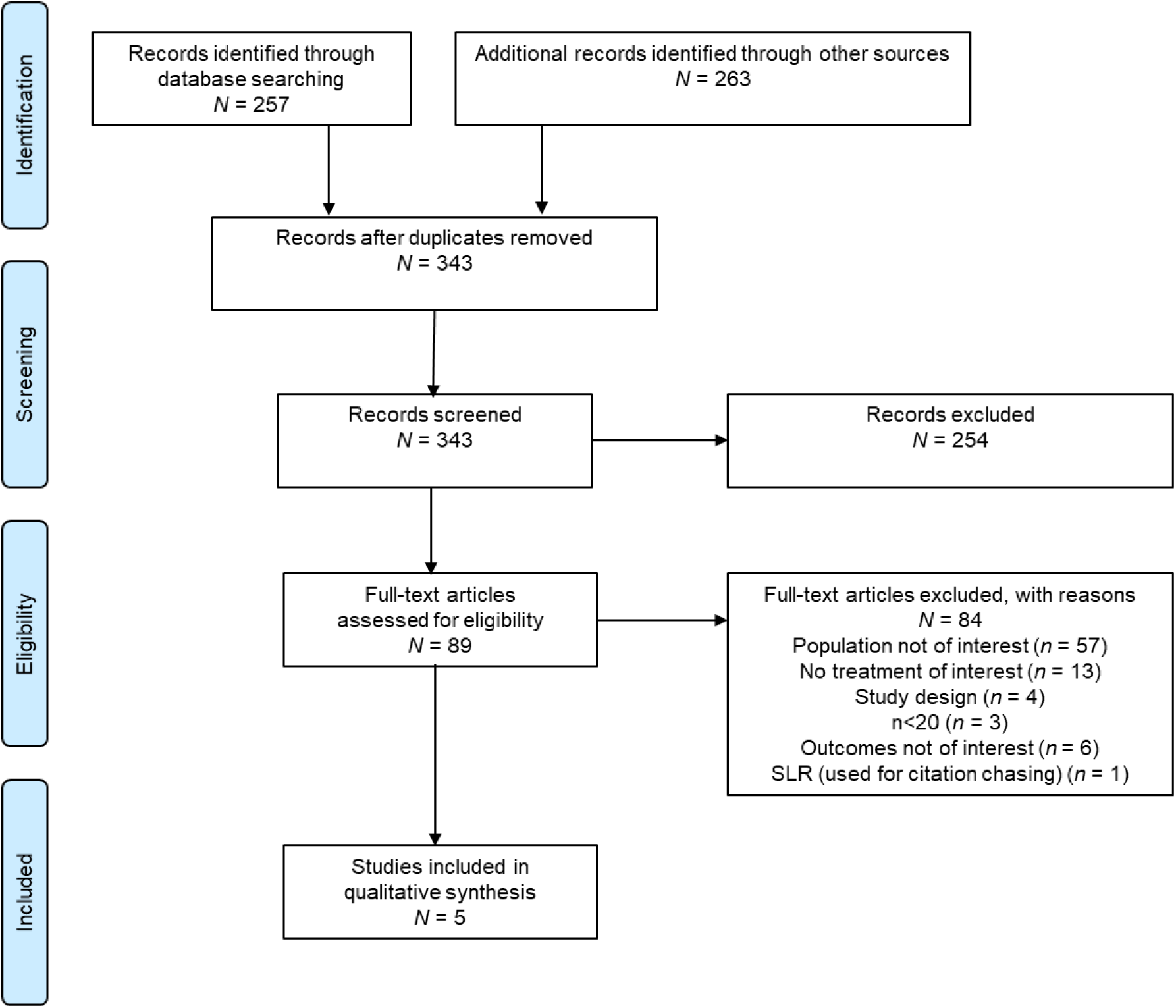
PRISMA flow diagram of studies included in the SLR. *PRISMA* Preferred Reporting Items for Systematic Reviews and Meta-Analyses, *SLR* systematic literature review

### Study characteristics

An overview of the key characteristics of the five studies included in the SLR is provided in Table 2. Of these studies, four were conducted by external investigators and one (Cheng et al.) was sponsored by GSK and Vir Biotechnology (note that some authors of Cheng et al. [MD and DCG] are also authors of this SLR) [25]. Studies were conducted in Italy (*n* = 1), Qatar (*n* = 1), England (*n* = 2), and the US (*n* = 1). Three studies employed an ecological design, with the date or month of COVID-19 diagnosis used as a proxy for the likelihood of an infection being attributable to the prevalent Omicron subvariant circulating in the country/region at the time [25, 28, 29]. The other two studies used sequencing data to ascertain the SARS-CoV-2 subvariant of infection [26, 27]. All studies included patients defined as being high-risk.

**Table 2.**
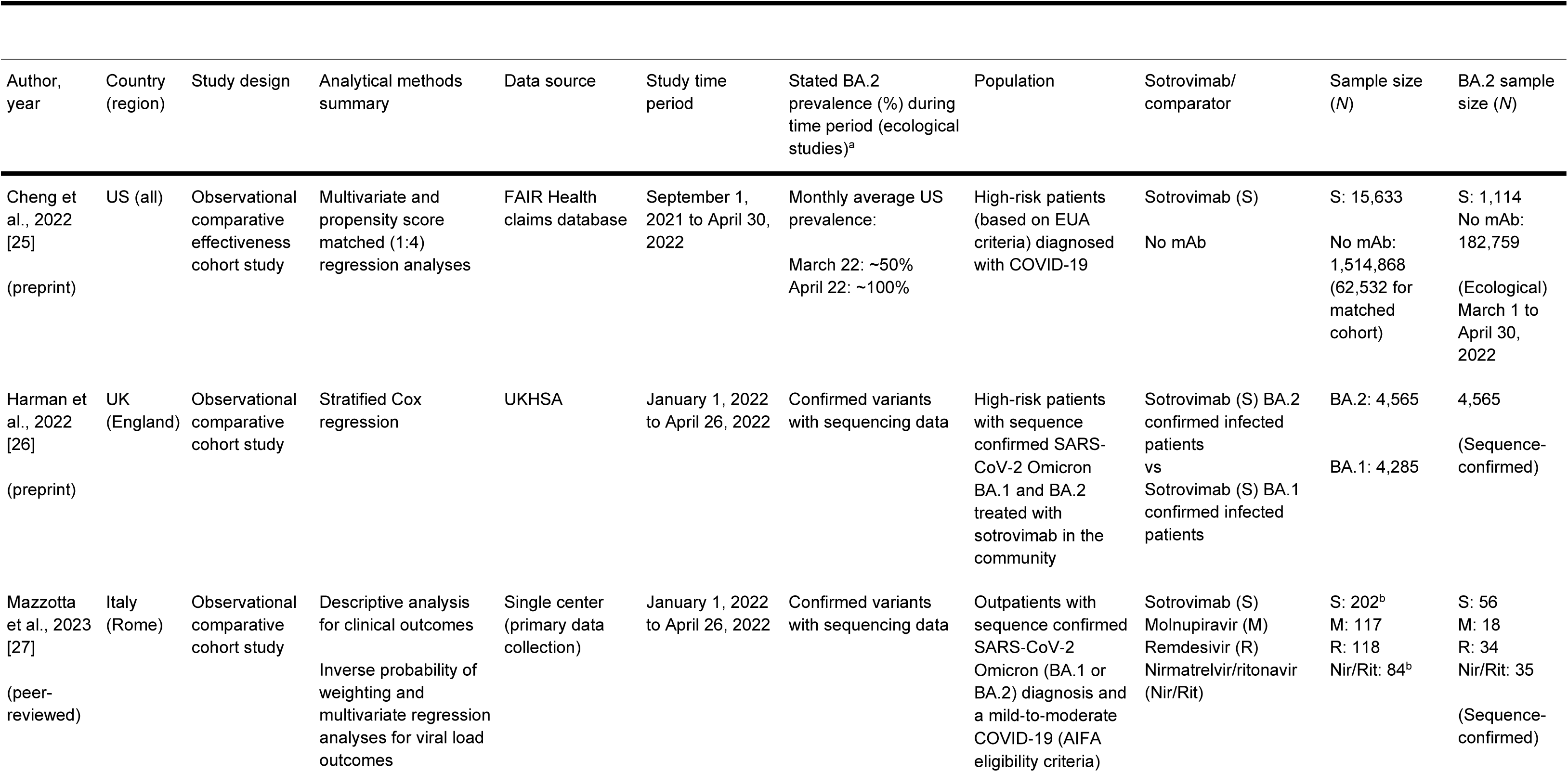
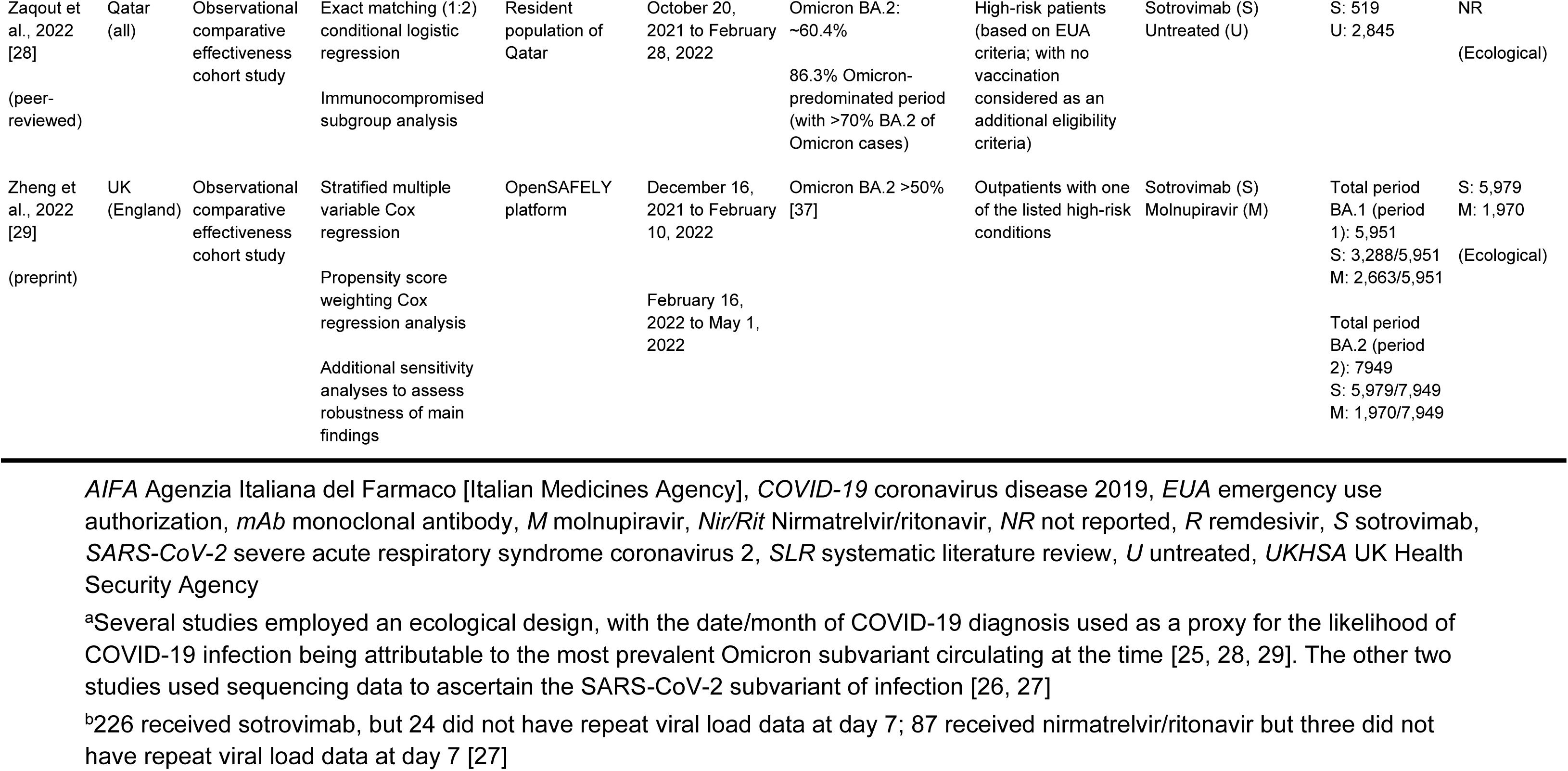
Overview of characteristics of studies included in the SLR

In total, these five studies included up to ∼1.5 million high-risk patients with COVID-19, of whom approximately 34,000 received sotrovimab as an early treatment for mild-to-moderate COVID-19 (approximately 12,000 of whom were treated during the period of Omicron BA.2 predominance). The high-risk populations included in the studies were heterogeneous, reflecting the differing treatment recommendations in each country at the time of study conduct. The population in the Cheng et al. study, conducted in the US [25], reflected the US EUA eligibility criteria for sotrovimab, as defined in the Infectious Diseases Society of America guidelines [30], which were very similar to the Agenzia Italiana del Farmaco guidelines [31] used in the Italian study by Mazzotta et al. [27]. Criteria such as an age of ≥65 (US) and >65 (Italy) years, or the presence of at least one comorbidity, such as obesity, diabetes, cardiovascular or chronic lung diseases, were not included in the NHS England guidelines for sotrovimab [32]. As NHS England had fewer criteria, the population eligible for receiving sotrovimab in the English studies by Harman et al. and Zheng et al. could be considered to be at an even higher risk [26, 29]. It should be noted that the two studies from England likely sampled from overlapping patient populations during the same time period. Finally, in Qatar, only 9% of residents are aged ≥50 years, which was reflected in the study population of Zaqout et al., and being unvaccinated was considered a risk factor, making the population less likely to match those identified as high-risk in other studies [28].

### Quality assessment

Out of the maximum attainable score of 9 on the NOS, three studies achieved a score of ≥7, suggesting that they were of comparatively good quality (Supplementary Table 2) [25, 26, 29]. The observational cohort studies by Cheng et al. in the US [25] and Zheng et al. in England [29] that used FairHealth claims data and the OpenSAFELY platform, respectively, were awarded a score of 8 and scored highly across all NOS domains. The observational cohort study by Harman et al. was awarded a score of 7 [26].

The remaining two studies were awarded a score of 6 [27, 28]. Mazzotta et al. was primarily designed to explore changes in SARS-CoV-2 viral load following treatment, and its score of 6 mainly reflects any shortcomings in assessing clinical outcomes rather than overall study quality. While viral load outcomes were adjusted for a range of clinical parameters, estimates of hospitalization and mortality were not [27]. Zaqout et al. was also awarded a score of 6 for some limitations in study design that may have introduced bias (Fig. 2) [28].

**Fig. 2.**
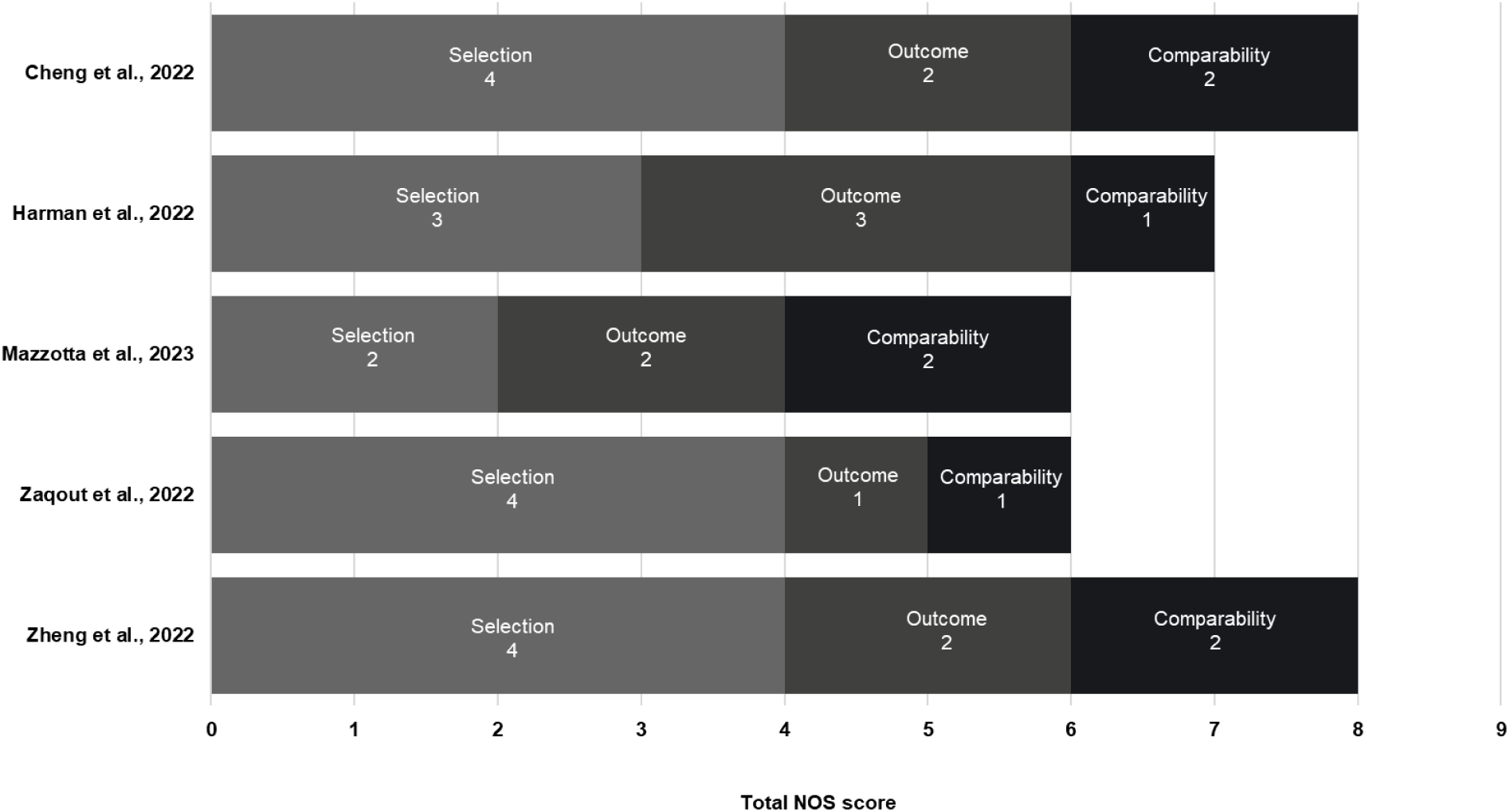
NOS total and bias domain scores across the studies included in the SLR. *NOS*, Newcastle Ottawa scale, *SLR* systematic literature review

It should be noted that NOS was used to assess the quality of each paper in its totality rather than by specific subgroups, endpoints, time periods, or SARS-CoV-2 variants. This is of particular relevance to the studies by Cheng et al. and Zaqout et al., which both included limited data on the Omicron BA.2 subvariant [25, 28]. The study by Cheng et al. was limited by the small sotrovimab sample size during March and April 2022 due to the deauthorization of sotrovimab, which led to wide confidence intervals for this period [25]. Due to staggered deauthorization of sotrovimab in the US at the time, this study was limited in its ability to assess the clinical effectiveness of sotrovimab during BA.2 predominance. The study by Zaqout et al. was also limited by its sample size during BA.2 predominance [28].

### Clinical outcomes

Of the five included studies, four reported on the composite measure of hospitalization or mortality, either related to COVID-19 [26, 27, 29] and/or all-cause [25, 26] during the period of Omicron BA.2 predominance. A single study, by Zheng et al., also reported estimates for mortality (due to any cause) alone [29]. Clinical outcomes were reported within 28 or 30 days of treatment, with the exception of Harman et al., which reported outcomes within 14 days of treatment [26]. Only one study (Zaqout et al., Qatar) described the results for progression to severe, critical, or fatal COVID-19 [28]. It should be noted that the reasons for COVID-related hospital admission in Qatar differed from other included studies; hospitalization was utilized as a means to proactively deploy treatment with the goal of preventing transmission and progression of COVID-19, as opposed to reducing the risk of further progression [33]. As such, any comparison of hospitalization proportions with the other studies should be undertaken with caution.

Four studies reported outcomes for sotrovimab during periods of both Omicron BA.1 and BA.2 predominance [25–27, 29]. Of note, Zaqout et al. only reported outcomes during a period when both Omicron BA.1 and BA.2 were circulating without differentiating outcomes by subvariant [28].

The clinical outcomes data extracted from the five studies included in this review are provided in Table 3. A summary of results deemed most pertinent to the objectives of this study, namely clinical outcomes during periods of Omicron predominance, when available, are presented in Fig. 3.

**Fig. 3.**
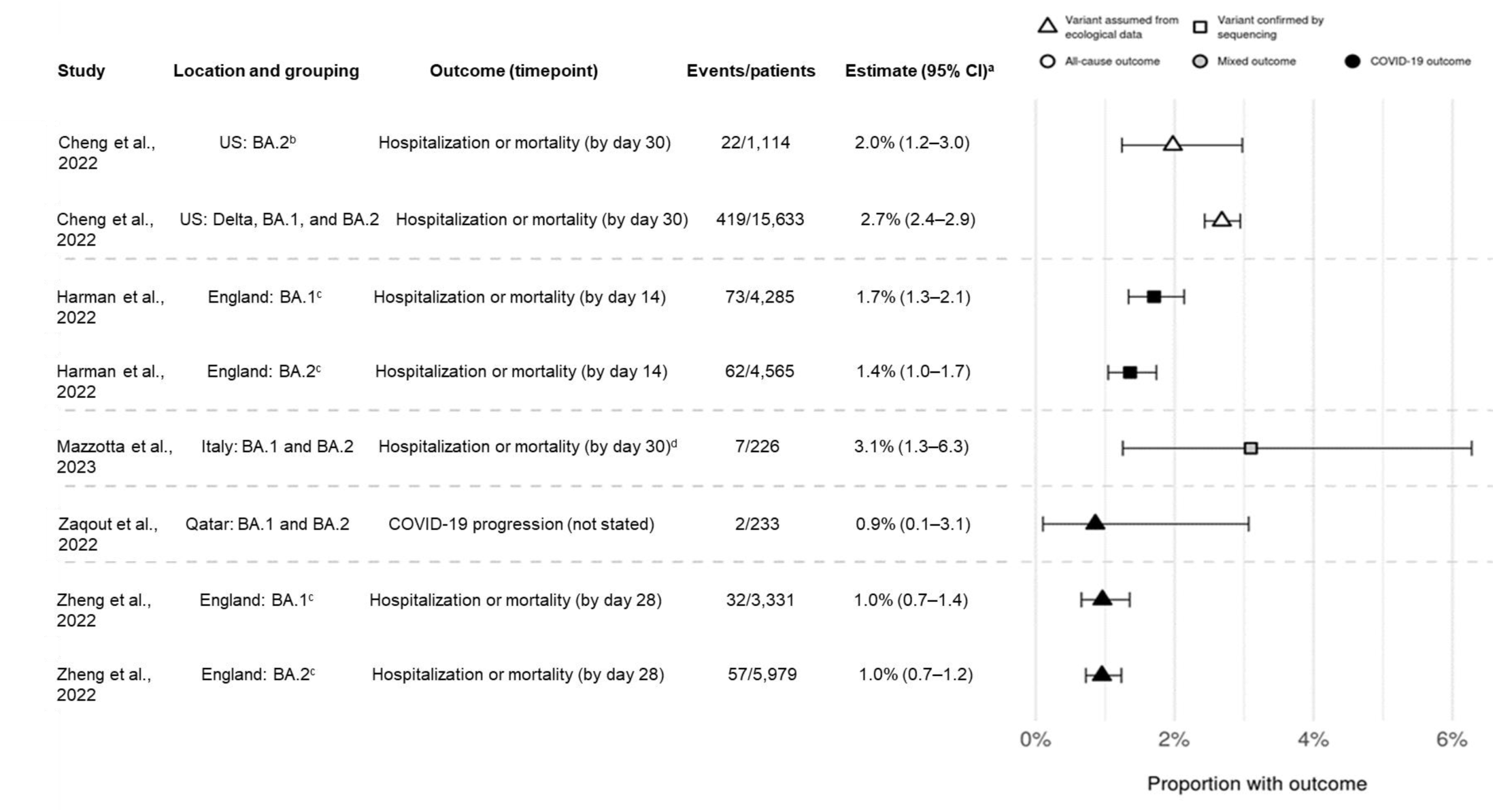
Point estimates for hospitalization or mortality (as a composite endpoint) or clinical progression for sotrovimab-treated patients. *^a^95 CIs calculated via Clopper-Pearson methods using reported data*. *^b^Defined as March through April 2022 in source and assumes homogeneity in the distribution of SARS-CoV-2 variants across all US states. ^c^Only COVID-19-specific outcome shown; all-cause outcome also reported in source. ^d^Hospitalizations were COVID-19-specific; deaths could be due to any cause*. *CI* confidence interval

**Table 3.** Clinical effectiveness of sotrovimab during Omicron BA.2 predominance

| Variant predominant | Outcome definition | Outcome time point | Sotrovimab (N) | Comparator (N) | Outcome N (%) |  | Relative effect (95% CI), significance |
| --- | --- | --- | --- | --- | --- | --- | --- |
|  |  |  |  |  | Sotrovimab | Comparator |  |
| Cheng et al., 2022 [25] |  |  |  |  |  |  |  |
| Overall (September 2021 through April 2022) | Hospitalization or mortality (all-cause) | 30 days of diagnosis | 15,633 | No mAb (unmatched: 1,514,868; matched: 62,532) | 419 (2.68) | Unmatched: 84,720 (5.59)<br>Matched: NR | RR 0.45 (0.41–0.49), $p < 0.0001^a$<br>PS-matched 0.39 (0.35–0.43), $p < 0.0001^b$ |
| March 2022 through April 2022 | Hospitalization or mortality (all-cause) | 30 days of diagnosis | March 2022: 1,046<br>April 2022: 68<br>Combined for BA.2: 1,114 | No mAb (unmatched March 2022: 65,521; April 2022: 117,238; combined for BA.2: 182,759; matched: NR) | March 2022: 21 (calculated, 2.01% of 1,046)<br>April 2022: 1 (calculated, 1.47% of 68)<br>Combined for BA.2: 22 (2.0) | March 2022: 2,863 (calculated, 4.37% of 65,521)<br>April 2022: 2,228 (calculated, 1.90% of 117,238)<br>Combined for BA.2: 5,091 (2.8)<br>Matched: NR | March 2022 RR 0.41 (0.27–0.62), $p < 0.0001^a$<br>March 2022 PS-matched 0.36 (0.23–0.56), $p < 0.0001^b$<br>April 2022 RR 0.54 (0.08–3.54), $p = 0.52^a$<br>April 2022 PS-Matched 0.32 (0.04–2.38), $p = 0.52^b$ |
| Harman et al., 2022 [26] |  |  |  |  |  |  |  |
| BA.2 vs BA.1 | Hospitalization or mortality (all-cause) | 14 days of treatment | BA.2 (4,565) | — | BA.2: 77 (1.7) | — | BA.2 vs BA.1<br>HR 1.17 (0.74—1.86), $p = \text{NR}^c$ |
|  |  |  | BA.1 (4,285) |  | BA.1: 91 (2.1) |  |  |
| BA.2 vs BA.1 | Hospitalization or mortality (COVID-19 related) | 14 days of treatment | BA.2 (4,565) | | BA.2: 62 (1.4) | | BA.2 vs BA.1<br>HR 0.98 (0.58—1.65), $p = \text{NR}^c$ |
|  |  |  | BA.1 (4,285) |  | BA.1: 73 (1.7) |  |  |
| Mazzotta et al., 2023 [27] |  |  |  |  |  |  |  |
| BA.1 | Hospitalization (COVID-19-related) or mortality (all-cause) | 30 days of treatment | 146 | Nirmatrelvir/Ritonavir (Nir/Rit) (49)<br>Remdesivir (R) (84)<br>Molnupiravir (M) (99) | 5<br>Overall<br>BA.1+BA.2: 7/226 (3.1) | Nir/Rit: 2<br>Overall<br>BA.1+BA.2: 2/87 (2.3)<br>R 0 (0)<br>M 0 (0) | NR |
| BA.2 | Hospitalization (COVID-19-related) or mortality (all-cause) | 30 days of treatment | 56 | Nir/Rit (35)<br>R (34)<br>M (18) | 2<br>Overall<br>BA.1+BA.2: 7/226 (3.1) | Nir/Rit: 0<br>Overall<br>BA.1+BA.2: 0/87<br>R 0 (0)<br>M 0 (0) | NR |
| Zaqout et al., 2022 [28] |  |  |  |  |  |  |  |
| Delta and Omicron | Progression to severe, critical, or fatal COVID-19 | NR | 345 | Untreated (583) | 4 (1.2) | 3 (0.5) | Adjusted OR 2.67 (0.60—11.91)<sup>d</sup> |
| Delta and Omicron | Progression to severe, critical, or fatal COVID-19 in patients at higher risk of severe COVID-19<sup>e</sup> | NR | 295 | Untreated (533) | 3 (1.0) | 8 (1.5) | Adjusted OR 0.65 (0.17–2.48)<sup>d</sup> |
| Omicron | Progression to severe, critical, or fatal COVID-19 | NR | 233 | Untreated (431) | 2 (0.9) | 0 (0) | NR |
| Omicron | Progression to severe, critical, or fatal COVID-19 in patients at higher risk of severe COVID-19<sup>e</sup> | NR | 210 | Untreated (391) | 2 (1.0) | 4 (1.0) | 0.88 (0.16–4.89)<sup>d</sup> |
| Zheng et al., 2022 [29] |  |  |  |  |  |  |  |
| BA.1 | Hospitalization or mortality (COVID-19-related) | 28 days of treatment | 3,331 | Molnupiravir (2,689) | 32 (0.96) | 55 (2.05) | Stratified Cox HR 0.54 (0.33–0.88), $p = 0.014^f$<br>PSW-Cox HR 0.50 (0.31–0.81), $p = 0.005^f$ |
| BA.2 | Hospitalization or mortality (COVID-19-related) | 28 days of treatment | 5,979 | Molnupiravir (1,970) | 57 (0.95) | 40 (2.03) | Stratified Cox HR 0.44 (0.27–0.71), $p = 0.001^f$<br>PSW-Cox HR 0.53 (0.32–0.86), $p = 0.010^f$ |
| BA.1 | Mortality<br>(COVID-19-related) | 28 days of<br>treatment | 3,331 | Molnupiravir<br>(2,689) | 7 (0.21) | 18 (0.67) | NR |
| BA.2 | Mortality<br>(COVID-19-related) | 28 days of<br>treatment | 5,979 | Molnupiravir<br>(1,970) | 9 (0.15) | 19 (0.96) | NR |

*CI* confidence interval, *COVID-19* coronavirus disease 2019, *HR* hazard ratio, *M* molnupiravir, *mAb* monoclonal antibody, *Nir/Rit* nirmatrelvir/ritonavir, *NR* not reported, *OR* odds ratio, *PS* propensity score, *PSW* propensity score weighted, *R* remdesivir, *RR* relative risk
<sup>a</sup>Adjusted for diagnosis month, category, age, gender, region, rurality, high-risk conditions, documented COVID-19 vaccine
<sup>b</sup>Matched on diagnosis month, age, gender, region, rurality, and selected high-risk conditions
<sup>c</sup>Hospitalization excluded hospital admissions for injury-related reasons. Adjusted for age group, linear effect in age and vaccination status to account for confounders
<sup>d</sup>Cases and controls were exact-matched one-to-two by vaccination status, prior infection status, sex, age group, nationality group, comorbidity count, and epidemic phase
<sup>e</sup>Defined as individuals who were immunocompromised (recipients of solid organ or hematopoietic stem cell transplant, patients receiving chemotherapy or immunosuppressive treatments, patients with severe immunodeficiency, and patients with human immunodeficiency virus ), unvaccinated individuals, those aged ≥75 years, and pregnant women
<sup>f</sup>Adjusted for age, sex, ten high-risk cohort categories, ethnicity, indices of multiple deprivation quintiles, vaccination status, calendar week, body mass index category, diabetes, hypertension, chronic cardiac and respiratory diseases

#### Descriptive clinical outcomes

The proportions of patients experiencing COVID-19-related hospitalization or mortality were consistently low across all studies and across periods of both Omicron BA.1 and BA.2 predominance. For sotrovimab-treated patients, COVID-19-related hospitalization or mortality ranged from 1.0% [29] to 3.1% [27] during Omicron BA.1 predominance, and from 1.0% [29] to 3.6% [27] during BA.2 predominance.

The proportions of patients experiencing all-cause hospitalization and mortality ranged between 2.1% and 2.7% for the Omicron BA.1 period, and 1.7% and 2.0% for the Omicron BA.2 period, as reported by Harman et al. (day 14) and Cheng et al. (day 30), respectively [25, 26]. Mortality as a standalone endpoint was only reported by Zheng et al.; COVID-19-related mortality was estimated at 0.21% (*n* = 7/3,331) for the sotrovimab group vs 0.67% (*n* = 18/2,689) for the molnupiravir group during Omicron BA.1 predominance, and 0.15% (*n* = 9/5,979) vs 0.96% (*n* = 19/1,970) during Omicron BA.2 predominance, respectively [29].

#### Clinical effectiveness of sotrovimab vs control/comparator

Three studies examined the clinical effectiveness of sotrovimab vs a control/comparator during the Omicron BA.2 predominance period [25, 28, 29].

The study by Zheng et al., which was conducted in England, demonstrated that sotrovimab was associated with a substantially lower risk of 28-day COVID-19-related hospitalization or mortality compared with molnupiravir during both the Omicron BA.1 and BA.2 subvariant surges [29]. Cox proportional hazards models indicated that after adjusting for demographics, high-risk cohort categories, vaccination status, calendar time, body mass index, and other comorbidities, sotrovimab was associated with a substantially lower risk of COVID-19-related hospitalization or death compared with molnupiravir during the Omicron BA.1 (adjusted hazard ratio (HR) 0.54, 95% CI 0.33–0.88; *p* = 0.014) and BA.2 (adjusted HR 0.44, 95% CI 0.27–0.71; *p* = 0.001) periods (Table 3).

The US-based study by Cheng et al. reported that sotrovimab was associated with a lower risk of 30-day all-cause hospitalization or mortality compared with no mAb treatment during the Omicron BA.2 subvariant surge in March and April 2022 (Table 3) [25]. In March 2022, sotrovimab effectiveness was significantly higher with an adjusted relative risk (RR) reduction of 59% (adjusted RR 0.41, 95% CI 0.27– 0.62) and a propensity score-matched RR reduction of 64% (adjusted RR 0.36, 95% CI 0.23–0.56) in 30-day all-cause hospitalization or mortality among sotrovimab-treated patients vs patients not treated with a mAb. In April 2022, the adjusted RR reduction in 30-day all-cause hospitalization or mortality among sotrovimab-treated patients was 46% (adjusted RR 0.54, 95% CI 0.08–3.54) and the propensity score-matched RR reduction was 68% (adjusted RR 0.32, 95% CI 0.04–2.38) compared with patients not treated with a mAb (Table 3). During Delta and Omicron BA.1 predominance (September 2021 to March 2022), treatment with sotrovimab compared with no mAb was associated with significant RR reductions in 30-day all-cause hospitalization or mortality ranging from 51% (December 2021, RR 0.49, 95% CI 0.43–0.57) to 71% (October 2021, RR 0.29, 95% CI 0.17–0.51) (propensity score-matched RR reductions from 55% (December 2021, RR 0.45, 95% CI 0.39–0.53) to 73% (October 2021, RR 0.27, 95% CI 0.15–0.47)) [25].

Zaqout et al. examined the real-world effectiveness of sotrovimab against COVID-19 in Qatar between October 20, 2021 and February 28, 2022 [28]. This study reported that the adjusted OR of disease progression to severe, critical, or fatal COVID-19 for sotrovimab vs the untreated control group over the entire study period was 2.67 (95% CI 0.60–11.91) (Table 3). Patients described as being at higher risk of severe forms of COVID-19 (immunocompromised, unvaccinated individuals, aged ≥75 years, and pregnant women), had lower odds of progression (adjusted OR 0.65, 95% CI 0.17–2.48).

When restricting the main analysis to the Omicron-predominant period (December 19, 2021 to February 28, 2022) an adjusted OR of disease progression could not be calculated, as none of the 431 untreated patients were observed to have progressed; two of the 233 (0.9%) patients treated with sotrovimab progressed during this phase (Table 3, Fig. 3). The analysis of the subgroup of patients at higher risk of severe forms of COVID-19 during this Omicron-predominated period yielded an adjusted OR of 0.88 (95% CI 0.16–4.89) (Table 3) [28].

Zaqout et al. described outcomes for study populations that they referred to as ‘main analysis’ and ‘subgroup analysis’. However, the ‘control’ cohorts for these two analyses were selected using different matching methodology; this approach is likely why a greater number of events were reported in the ‘subgroup analysis’ control group than that observed in the ‘main analysis’ control group.

#### Clinical outcomes with sotrovimab in treating Omicron BA.1 vs BA.2

A single study, conducted by Harman et al. in England, directly compared clinical outcomes of sotrovimab-treated patients infected with Omicron BA.1 (*n* = 4,285) vs Omicron BA.2 (*n* = 4,565), as confirmed by sequencing [26]. The results of this study suggested that the risk of hospital admission was similar between Omicron BA.1 and BA.2 infections treated with sotrovimab (Table 3); there was no evidence of a difference in the risk of hospital admission with a length of stay of ≥2 days within 14 days of sotrovimab treatment between the BA.1 (2.1%, *n* = 91) and BA.2 (1.7%, *n* = 77) subvariants (HR 1.17, 95% CI 0.74–1.86) [26].

## Discussion

This SLR identified and assessed all observational studies in the published literature available as of November 3, 2022, which reported clinical outcomes for patients treated with sotrovimab during Omicron BA.2 subvariant predominance and onwards circulating variants. In this context, real-world evidence is potentially a more agile source of evidence than randomized clinical trials.

A recently published SLR and meta-analysis by Amani et al. demonstrated the real-world effectiveness of sotrovimab in terms of reducing hospitalization and mortality during both the Delta and Omicron BA.1 periods of predominance [20]. The findings of the current SLR build on the work of Amani et al. and demonstrate the real-world benefit of sotrovimab for the treatment of COVID-19 during the Omicron BA.2 predominance period. The studies included in our review consistently reported low proportions of severe clinical outcomes (such as all-cause or COVID-19-related hospitalization or mortality) in patients treated with sotrovimab during the predominant period of Omicron BA.2. In addition, although only a limited number of studies evaluated the clinical outcomes of sotrovimab during both the Omicron BA.1 and BA.2 periods, these demonstrated that clinical outcomes in patients with COVID-19 treated with sotrovimab were consistently low across Omicron BA.1 and BA.2 predominance periods. Furthermore, one large study by Harman et al. found no evidence of a difference in clinical outcomes when directly comparing patients treated with sotrovimab with sequencing-confirmed BA.1 and BA.2 [26]. Together, these findings provide no evidence to indicate that the neutralization fold change reported in vitro led to a commensurate change in the effectiveness of sotrovimab.

The low proportions of severe clinical outcomes summarized in the current SLR closely align with the 1% all-cause hospitalization or mortality through day 29 reported for sotrovimab in the randomized COMET-ICE trial conducted when the wild-type strain was predominant [13]. These real-world clinical effectiveness data were generated from recent use of sotrovimab in patient populations as recommended by country-specific guidelines, and hence reflect the clinical risk and immunological characteristics of the patient population more closely than clinical trials. In particular, population-level immunity resulting from both vaccination and prior infection means these effectiveness results provide important information for prescribers, as the COMET-ICE population was unvaccinated and likely immunologically naïve.

In the current SLR, two high-quality studies from England were included [26, 29]. The observational cohort study by Zheng et al. leveraged the substantial size of the OpenSAFELY platform database to examine the effectiveness of sotrovimab in preventing severe COVID-19 outcomes across both the Omicron BA.1 and BA.2 periods of predominance using propensity scoring methodology and a number of sensitivity analyses to confirm the robustness of the analyses [29]. This study demonstrated that sotrovimab was associated with a substantially lower risk of 28-day COVID-19-related hospitalization or mortality during the Omicron BA.2 subvariant surge compared with molnupiravir after adjustment. The proportions of COVID-19-related hospitalization or mortality for sotrovimab were also comparable across Omicron BA.1 and BA.2. Lower mortality in patients treated with sotrovimab vs molnupiravir was also reported during both Omicron periods of predominance. Zheng et al. concluded that these data support a persistent protective role for sotrovimab against the Omicron BA.2 subvariant [29]. It should be noted, however, that guidance in England for molnupiravir was changed from a second- to third-line treatment option between the Omicron BA.1 and BA.2 periods of predominance, while sotrovimab remained a first-line option during both periods [32]. Although the impact of this change in national recommendations is unclear, it may have altered the baseline characteristics of patients who received molnupiravir in the Zheng et al. study, and the analysis of the BA.2 period was considered exploratory by the authors. Multiple sensitivity analyses were undertaken as part of this study, and the consistency of the results was maintained.

The results from Zheng et al. are supported by Harman et al. [26]. This large retrospective cohort study of SARS-CoV-2-sequenced patients in England assessed the risk of hospital admission or mortality within 14 days in patients treated with sotrovimab and infected with Omicron BA.2, compared with Omicron BA.1. No evidence of a difference between the Omicron BA.2 and BA.1 subvariants was observed. However, it should be noted that testing guidance in England varied during Omicron predominance, and free community testing was restricted from April 1, 2022. This reduced sequencing capacity and thus impacted the overall number of cases available for inclusion in Harman et al.; possible selection bias may have been introduced after this date as a result. In addition, the absence of a comparator-treated control group, and the limited information on comorbidities and severity, limit the utility of the study in assessing the effectiveness of sotrovimab during the Omicron BA.2 period. Nevertheless, the fact that the results of both the Zheng et al. and Harman et al. studies are consistent across different clinical outcomes further supports the robustness of these findings. In addition, the findings of the ecological study conducted by Zheng et al. are aligned with the findings of Harman et al., where variant of infection was confirmed by sequencing. The remainder of the studies identified in the SLR are consistent in reporting low rates of severe clinical outcomes in sotrovimab-treated patients during periods of Omicron BA.1 and BA.2 predominance.

A single study from Zaqout et al., however, reported a point estimate for the main finding of progression to severe, critical, or fatal COVID-19 in favor of the comparator untreated group [28]. These results had wide confidence intervals and were non-significant, and it is notable that the point estimate is favorable for sotrovimab when the analysis population is limited to those only at higher risk. It should be noted that a selection bias toward patients less likely to progress to severe disease was expected for the control group in this point estimate, as patients were excluded from the control group if they showed signs or symptoms of severe COVID-19 within 7 days of diagnosis.

Two additional studies that did not meet the inclusion criteria of this SLR but support its findings (consistent clinical benefit with sotrovimab during the Omicron BA.2 subvariant predominant period) were identified. Interim results of the French multicenter, prospective, observational cohort study, COCOPREV, were published as a Letter to the Editor at the time of the review and were, therefore, out of scope [34]. These results indicated low and similar proportions of hospitalization or mortality within 28 days of sotrovimab treatment in patients infected with Omicron BA.1 (*n* = 125; 2.4%; 95% CI 1–7) and BA.2 (*n* = 42; 2.4%; 95% CI 0–13) viral variants, as confirmed by sequencing. No patients died in either group. In addition, there was no evidence of a difference in the slope of the change over time in the cycle threshold values between Omicron BA.1 or BA.2 infected patients (*p* = 0.87), indicating that time to virus resolution was similar between the two groups. It should be noted that the sample size of Omicron BA.2 infected patients in COCOPREV was comparatively small [34]. Secondly, the results of an interim report of a Japanese post-marketing study were only published in Japanese at the time our SLR was conducted and were thus excluded. Results were subsequently published in English and demonstrate a similarity in clinical outcomes for sotrovimab-treated patients infected with both Omicron BA.1 and BA.2 [35]. Progression (defined as needing oxygen or ventilation, needing ICU for exacerbation, hospitalization for exacerbation, or death due to exacerbation) within 29 days of sotrovimab administration or discharge/transfer date was assessed in hospitalized patients with mild-to-moderate COVID-19 (*n* = 246 for clinical outcomes). The rate of progression was found to be similar between the groups: 0.8% (95% CI 0.02–4.63) in Omicron BA.1 (January 31, 2022 to March 27, 2022) and 0% (95% CI 0.00–2.84) during BA.2 (March 28, 2022 to June 19, 2022). While many patient characteristics were similar across the periods, small differences in sex, age, weight, comorbidity status, vaccination status, and body temperature were reported, and not corrected for. It should also be noted that hospitalization in Japan was not only for clinical reasons, which may have affected these findings [35].

### Limitations

This SLR has several limitations that should be considered. Firstly, the number of studies identified in this SLR is small, although they collectively included a large number of participants. Due to the rapidly evolving landscape around COVID-19, real-world data for sotrovimab are still emerging, and it is expected that additional observational studies will further contribute to the understanding of sotrovimab’s effectiveness during the recent period of Omicron BA.2 predominance. Secondly, two studies published in preprint databases have been included in this SLR [25, 26]. While these should be interpreted with caution, as they are not peer-reviewed, preprint publication has been commonly used throughout the COVID-19 pandemic to rapidly report outcomes so as to guide responsive decision-making around urgent public health matters [36]. In addition, due to a lack of sequencing data, several studies used an ecological design to infer the causative variant using the date of SARS-CoV-2 infection [25, 28, 29]. Mazzotta et al. and Harman et al. used sequencing data to fully ascertain the SARS-CoV-2 subvariant of infection [26, 27]. Finally, a meta-analysis was not considered feasible as the included studies were diverse in terms of population of interest, target outcomes, study design, and analytical methods applied to estimate clinical outcomes during Omicron BA.2; combining studies may amplify the presence of confounding factors.

## Conclusions

Results from this SLR suggest continued clinical effectiveness of sotrovimab (IV 500 mg) in preventing severe clinical outcomes related to COVID-19 infections during the period of Omicron BA.2 predominance vs control/comparator and compared with the period of Omicron BA.1 predominance, despite reduced in vitro neutralization activity. The studies included in this review were consistent in reporting low proportions of severe clinical outcomes (such as hospitalization and mortality) in sotrovimab-treated patients during the periods of Omicron BA.1 and Omicron BA.2 subvariant predominance.

## Declarations

### Author contribution

MD, AS and EJL were involved in the conception of this study. MD, DCG, CR and LL performed the literature review, and all authors were involved in the data analysis. All authors took part in drafting, revising or critically reviewing the manuscript; gave final approval of the version to be published; have agreed on the journal to which the article has been submitted; and agree to be accountable for all aspects of the work in ensuring that questions related to the accuracy or integrity of any part of the work are appropriately investigated and resolved.

### Data availability statement

All datasets generated for this study are included in this manuscript.

### Ethical approval

Only publicly available papers were included in this SLR, and no human subjects were involved; ethics approval was therefore not required.

### Informed consent

Not applicable.

### Consent to participate

Not applicable. Consent to publish Not applicable.

## Supporting information

219682 SLR pre-print supplement

## Data Availability

All datasets generated for this study are included in this manuscript.

## Acknowledgments

Editorial support (in the form of writing assistance, including preparation of the draft manuscript under the direction and guidance of the authors, collating and incorporating authors’ comments for each draft, assembling tables and figures, grammatical editing, and referencing) was provided by Kathryn Wardle of Aura, a division of Spirit Medical Communications Group Limited (Manchester, UK), and was funded by GSK and Vir.

## Supplementary material

Search strategy and quality assessment tables

## References

1. Cucinotta D, Vanelli M. WHO declares COVID-19 a pandemic. Acta Biomed. 2020;91:157–60. doi:10.23750/abm.v91i1.9397

2. Dessie ZG, Zewotir T. Mortality-related risk factors of COVID-19: a systematic review and meta-analysis of 42 studies and 423,117 patients. BMC Infect Dis. 2021;21:855. doi:10.1186/s12879-021-06536-3

3. Hippisley-Cox J, Khunti K, Sheikh A, Nguyen-Van-Tam JS, Coupland CAC. QCovid 4 - Predicting risk of death or hospitalisation from COVID-19 in adults testing positive for SARS-CoV-2 infection during the Omicron wave in England. medRxiv. 2022:2022.08.13.22278733. doi:10.1101/2022.08.13.22278733

4. World Health Organization. Coronavirus (COVID-19) dashboard. 2022; https://covid19.who.int/. Accessed 22 Dec 2022.

5. Shadmi E, Chen Y, Dourado I, Faran-Perach I, Furler J, Hangoma P, et al. Health equity and COVID-19: global perspectives. Int J Equity Health. 2020;19:104. doi:10.1186/s12939-020-01218-z

6. Chaudhry R, Dranitsaris G, Mubashir T, Bartoszko J, S R. A country level analysis measuring the impact of government actions, country preparedness and socioeconomic factors on COVID-19 mortality and related health outcomes. EClinicalMedicine. 2020;25:100464.

7. Mendiola-Pastrana IR, López-Ortiz E, Río de la Loza-Zamora JG, González J, Gómez-García A, López-Ortiz G. SARS-CoV-2 variants and clinical outcomes: a systematic review. Life (Basel). 2022;12:170. doi:10.3390/life12020170

8. Gaudinski MR, Coates EE, Houser KV, Chen GL, Yamshchikov G, Saunders JG, et al. Safety and pharmacokinetics of the Fc-modified HIV-1 human monoclonal antibody VRC01LS: a phase 1 open-label clinical trial in healthy adults. PLoS Med. 2018;15:e1002493. doi:10.1371/journal.pmed.1002493

9. Ko SY, Pegu A, Rudicell RS, Yang ZY, Joyce MG, Chen X, et al. Enhanced neonatal Fc receptor function improves protection against primate SHIV infection. Nature. 2014;514:642–5. doi:10.1038/nature13612

10. Cathcart AL, Havenar-Daughton C, Lempp FA, Ma D, Schmid M, Agostini ML, et al. The dual function monoclonal antibodies VIR-7831 and VIR-7832 demonstrate potent in vitro and in vivo activity against SARS-CoV-2. bioRxiv. 2021:2021.03.09.434607. doi:10.1101/2021.03.09.434607

11. Pinto D, Park Y-J, Beltramello M, Walls AC, Tortorici MA, Bianchi S, et al. Cross-neutralization of SARS-CoV-2 by a human monoclonal SARS-CoV antibody. Nature. 2020;583:290–5. doi:10.1038/s41586-020-2349-y

12. Gupta A, Gonzalez-Rojas Y, Juarez E, Crespo Casal M, Moya J, Falci DR, et al. Early treatment for covid-19 with SARS-CoV-2 neutralizing antibody sotrovimab. N Engl J Med. 2021;385:1941–50. doi:10.1056/NEJMoa2107934

13. Gupta A, Gonzalez-Rojas Y, Juarez E, Crespo Casal M, Moya J, Rodrigues Falci D, et al. Effect of sotrovimab on hospitalization or death among high-risk patients with mild to moderate COVID-19: a randomized clinical trial. JAMA. 2022;327:1236–46. doi:10.1001/jama.2022.2832

14. GSK and Vir Biotechnology announce sotrovimab (VIR-7831) receives emergency use authorization from the US FDA for treatment of mild-to-moderate COVID-19 in high-risk adults and paediatric patients [press release]. 26 May 2021.

15. European Medicines Agency. Xevudy. 2023; https://www.ema.europa.eu/en/medicines/human/EPAR/xevudy. Accessed 3 Feb 2023.

16. World Health Organization. Tracking SARS-CoV-2 variants. 2023; https://www.who.int/activities/tracking-SARS-CoV-2-variants. Accessed 18 Jan 2023.

17. World Health Organization. Weekly epidemiological update on COVID-19 - 22 March 2022. 2022; https://www.who.int/publications/m/item/weekly-epidemiological-update-on-covid-19---22-march-2022. Accessed 22 Dec 2022.

18. Park YJ, Pinto D, Walls AC, Liu Z, De Marco A, Benigni F, et al. Imprinted antibody responses against SARS-CoV-2 omicron sublineages. Science. 2022;378:619–27. doi:10.1126/science.adc9127

19. United States Food and Drug Administration. FDA updates sotrovimab emergency use authorization. 2022; https://www.fda.gov/drugs/drug-safety-and-availability/fda-updates-sotrovimab-emergency-use-authorization. Accessed 22 Dec 2022.

20. Amani B, Amani B. Efficacy and safety of sotrovimab in patients with COVID-19: a rapid review and meta-analysis. Rev Med Virol. 2022;32:e2402. doi:10.1002/rmv.2402

21. Page MJ, McKenzie JE, Bossuyt PM, Boutron I, Hoffmann TC, Mulrow CD, et al. The PRISMA 2020 statement: an updated guideline for reporting systematic reviews. BMJ. 2021;372:n71. doi:10.1136/bmj.n71

22. Higgins JP, Green S. Cochrane handbook for systematic reviews of interventions. Vol 4: John Wiley & Sons; 2011.

23. Ottawa Hospital Research Institute. The Newcastle-Ottawa Scale (NOS) for assessing the quality of nonrandomised studies in meta-analyses. 2021; https://www.ohri.ca/programs/clinical_epidemiology/oxford.asp. Accessed 22 Dec 2022.

24. Sanderson S, Tatt ID, Higgins JP. Tools for assessing quality and susceptibility to bias in observational studies in epidemiology: a systematic review and annotated bibliography. Int J Epidemiol. 2007;36:666–76. doi:10.1093/ije/dym018

25. Cheng MM, Reyes C, Satram S, Birch H, Gibbons DC, Drysdale M, et al. Real-world effectiveness of sotrovimab for the early treatment of COVID-19 during SARS-CoV-2 delta and omicron waves in the United States. medRxiv. 2022:2022.09.07.22279497. doi:10.1101/2022.09.07.22279497

26. Harman K, Nash SG, Webster HH, Groves N, Hardstaff J, Bridgen J, et al. Comparison of the risk of hospitalisation among BA.1 and BA.2 COVID-19 cases treated with sotrovimab in the community in England. medRxiv. 2022:2022.10.21.22281171. doi:10.1101/2022.10.21.22281171

27. Mazzotta V, Cozzi Lepri A, Colavita F, Rosati S, Lalle E, Cimaglia C, et al. Viral load decrease in SARS-CoV-2 BA.1 and BA.2 omicron sublineages infection after treatment with monoclonal antibodies and direct antiviral agents. J Med Virol. 2023:e28186. doi:10.1002/jmv.28186

28. Zaqout A, Almaslamani MA, Chemaitelly H, Hashim SA, Ittaman A, Alimam A, et al. Effectiveness of the neutralizing antibody sotrovimab among high-risk patients with mild-to-moderate SARS-CoV-2 in Qatar. Int J Infect Dis. 2022;124:96–103. doi:10.1016/j.ijid.2022.09.023

29. Zheng B, Green ACA, Tazare J, Curtis HJ, Fisher L, Nab L, et al. Comparative effectiveness of sotrovimab and molnupiravir for prevention of severe covid-19 outcomes in patients in the community: observational cohort study with the OpenSAFELY platform. BMJ. 2022;379:e071932. doi:10.1136/bmj-2022-071932

30. Bhuimraj A, Morgan RL, Hirsch Shumaker A, Baden L. Infectious Diseases Society of America guidelines on the treatment and management of patients with COVID-19. 2022.

31. Agenzia Italiana del Farmaco. AIFA recommendations on medicines to be used in home management of COVID-19 cases. 2022; https://www.aifa.gov.it/documents/20142/1269602/EN_Raccomandazioni_AIFA_gestione_domiciliare_COVID-19_Vers9_31.05.2022.pdf. Accessed 18 Jan 2023.

32. NHS England. Interim Clinical Commissioning Policy: treatments for hospital-onset COVID-19. 2022; https://www.england.nhs.uk/coronavirus/publication/interim-clinical-commissioning-policy-antivirals-or-neutralising-monoclonal-antibodies-in-the-treatment-of-hospital-onset-covid-19/. Accessed 22 Dec 2022.

33. Ministry of Public Health – State of Qatar. Interim guidelines for management of suspected/ confirmed cases of coronavirus. 2022; https://covid19.moph.gov.qa/EN/Pages/default.aspx. Accessed 22 Dec 2022.

34. Martin-Blondel G, Marcelin AG, Soulié C, Kaisaridi S, Lusivika-Nzinga C, Dorival C, et al. Sotrovimab to prevent severe COVID-19 in high-risk patients infected with omicron BA.2. J Infect. 2022;85:e104–8. doi:10.1016/j.jinf.2022.06.033

35. Nose Y, Yamamoto M, Mizohata H, Kaneuchi Y, Oda K, Handa Y, et al. Evaluation of safety and clinical outcomes of sotrovimab in patients infected with SARS-CoV-2 in real world clinical practice. Ther Res. 2022;43:795–815.

36. Watson C. Rise of the preprint: how rapid data sharing during COVID-19 has changed science forever. Nature Medicine. 2022;28:2–5. doi:10.1038/s41591-021-01654-6

37. UK Health Security Agency. SARS-CoV-2 variants of concern and variants under investigation in England: technical briefing 43. 2022; https://assets.publishing.service.gov.uk/government/uploads/system/uploads/attachment_data/file/1103533/Technical-Briefing-43-24June2022.pdf. Accessed 22 Dec 2022.

