## Supplementary material for "Real-world effectiveness of sotrovimab for the treatment of SARS-CoV-2 infection during Omicron BA.2 subvariant predominance: a systematic literature review": 219682 SLR pre-print supplement

| **Supplementary Table 1** Embase search strategy | |
| --- | --- |
| Search number | Search terms |
| 1 | coronavirus disease 2019/ or (covid-19 or covid19 or corona-virus or sars-cov-2 or sars-cov2 or coronavirus disease or ncov or n-cov).ti,ab. or (alpha or beta or gamma or delta or omicron).ti. |
| 2 | sotrovimab/ or (sotrovimab or GSK-4182136 or GSK4182136 or UNII-1MTK0BPN8V or VIR-7831 or VIR7831 or Xevudy).ti,ab. |
| 3 | 1 and 2. |
| 4 | exp Clinical trial/ or exp Randomized controlled trial/ or Randomization/ or Single blind procedure/ or Double blind procedure/ or Crossover procedure/ or Placebo/ or Randomi?ed controlled trial$.tw. or Rct.tw. or Random allocation.tw. or Randomly allocated.tw. or Allocated randomly.tw. or (allocated adj2 random).tw. or Single blind$.tw. or Double blind$.tw. or ((treble or triple) adj blind$).tw. or Placebo$.tw. |
| 5 | exp longitudinal study/ or exp retrospective study/ or exp prospective study/ or exp cohort analysis/ or exp cross-sectional study/ or exp cohort analysis/ or exp observational study/ or (longitudinal study or retrospective study or prospective study or cohort$ or follow up or cross-sectional study or cross sectional study or followup study or observational study or registry or registries or real world or cross sectional or claims database or electronic health record$ or EHR or electronic medical record$ or EMR$ or RWE).ti,ab. |
| 6 | meta-analysis/ or systematic review/ or meta-analysis as topic/ or "meta analysis (topic)"/ or "systematic review (topic)"/ or exp technology assessment, biomedical/. |
| 7 | ((systematic* adj3 (review* or overview*)) or (methodologic* adj3 (review* or overview*))).ti,ab. |
| 8 | ((quantitative adj3 (review* or overview* or synthes*)) or (research adj3 (integrati* or overview*))).ti,ab. |
| 9 | ((integrative adj3 (review* or overview*)) or (collaborative adj3 (review* or overview*)) or (pool* adj3 analy*)).ti,ab. |
| 10 | (data synthes* or data extraction* or data abstraction*).ti,ab. |
| 11 | (handsearch* or hand search*).ti,ab. |
| 12 | (mantel haenszel or peto or der simonian or dersimonian or fixed effect* or latin square*).ti,ab. |
| 13 | (meta analy* or metanaly* or technology assessment* or HTA or HTAs or technology overview* or technology appraisal*).ti,ab. |
| 14 | (meta regression* or metaregression*).ti,ab. |
| 15 | (meta-analy* or metaanaly* or systematic review* or biomedical technology assessment* or bio-medical technology assessment*).mp,hw. |
| 16 | (medline or cochrane or pubmed or medlars or embase or cinahl).ti,ab,hw. |
| 17 | (cochrane or (health adj2 technology assessment) or evidence report).jw. |
| 18 | (comparative adj3 (efficacy or effectiveness)).ti,ab. |
| 19 | (outcomes research or relative effectiveness).ti,ab. |
| 20 | ((indirect or indirect treatment or mixed-treatment) adj comparison*).ti,ab. |
| 21 | or/4-20. |
| 22 | 3 and 21. |
| 23 | limit 22 to (conference abstract and yr="2022-current"). |
| 24 | limit 22 to (yr="2022-current" and (article or article in press)). |
| 25 | 23 or 24. |

| **Supplementary Table 2** NOS cohort studies quality assessment of studies included in the SLR | | | | | | | |
| --- | --- | --- | --- | --- | --- | --- | --- |
| Bias domain | Questions | Response options (* = response scores a point) | Cheng et al. 2022 [1] | Harman et al. 2022 [2] | Mazzotta et al. 2023 [3] | Zaqout et al. 2022 [4] | Zheng et al. 2022 [5] |
| Selection | 1. Representativeness of the exposed cohort | a) Truly representative of the average high-risk COVID-19 patient in the community* | ✓ |  |  |  | ✓ |
|  |  | b) Somewhat representative of the average high-risk COVID-19 patient in the community* |  | ✓ | ✓ | ✓ |  |
|  |  | c) Selected group of users (e.g. nurses, volunteers) |  |  |  |  |  |
|  |  | d) No description of the derivation of the cohort |  |  |  |  |  |
|  | 2. Selection of non-exposed cohort | a) Drawn from the same community as the exposed cohort* | ✓ | ✓ | ✓ | ✓ | ✓ |
|  |  | b) Drawn from a different source |  |  |  |  |  |
|  |  | c) No description of the derivation of the non-exposed cohort |  |  |  |  |  |
|  | 3. Ascertainment of exposure | a) Secure record (e.g. surgical records)* | ✓ | ✓ |  | ✓ | ✓ |
|  |  | b) Structured interview* |  |  |  |  |  |
|  |  | c) Written self-report |  |  |  |  |  |
|  |  | d) No description |  |  | ✓ |  |  |
|  | 4. Demonstration that outcome of interest was not present at start of study | a) Yes* | ✓ |  |  | ✓ | ✓ |
|  |  | b) No |  | ✓ | ✓ |  |  |
| Comparability | 1. Comparability of cohorts on the basis of the design or analysis | a) Study controls for immunodeficiency (by disease and/or treatment)* | ✓ |  | ✓ |  | ✓ |
|  |  | b) Study controls for any additional factor* | ✓ | ✓ | ✓ | ✓ | ✓ |
| Outcome | 1. Assessment of outcome | a) Independent blind assessment* |  |  |  |  |  |
|  |  | b) Record linkage* | ✓ | ✓ |  | ✓ | ✓ |
|  |  | c) Self-report |  |  |  |  |  |
|  |  | d) No description |  |  | ✓ |  |  |
|  | 2. Was follow-up long enough for outcomes to occur | a) Yes* | ✓ | ✓ | ✓ |  | ✓ |
|  |  | b) No |  |  |  | ✓ |  |
|  | 3. Adequacy of follow-up of cohorts | a) Complete follow up – all subjects accounted for* |  | ✓ |  |  |  |
|  |  | b) Subjects lost to follow up unlikely to introduce bias – small number lost – >80% follow up, or description provided of those lost* |  |  | ✓ |  |  |
|  |  | c) Follow up rate <80% (select an adequate %) and no description of those lost |  |  |  |  |  |
|  |  | d) No statement | ✓ |  |  | ✓ | ✓ |
| Total score (max. 9) | | | 8 | 7 | 6 | 6 | 8 |

*NOS* Newcastle Ottawa Scale, *SLR* systematic literature review

Ticked boxes represent a score of 1, while empty boxes represent a score of 0

**Supplementary references**

1. Cheng MM, Reyes C, Satram S, Birch H, Gibbons DC, Drysdale M, et al. Real-world effectiveness of sotrovimab for the early treatment of COVID-19 during SARS-CoV-2 Delta and Omicron waves in the United States. medRxiv. 2022:2022.2009.2007.22279497. Preprint at https://doi.org/10.1101/2022.09.07.22279497 (2022).
2. Harman K, Nash SG, Webster HH, Groves N, Hardstaff J, Bridgen J, et al. Comparison of the risk of hospitalisation among BA.1 and BA.2 COVID-19 cases treated with sotrovimab in the community in England. medRxiv. 2022:2022.2010.2021.22281171. Preprint at https://doi.org/10.1101/2022.10.21.22281171 (2022).
3. Mazzotta V, Cozzi Lepri A, Colavita F, Rosati S, Lalle E, Cimaglia C, et al. Viral load decrease in SARS-CoV-2 BA.1 and BA.2 Omicron sublineages infection after treatment with monoclonal antibodies and direct antiviral agents. J Med Virol. 2023:e28186. https://doi.org/10.1002/jmv.28186.
4. Zaqout A, Almaslamani MA, Chemaitelly H, Hashim SA, Ittaman A, Alimam A, et al. Effectiveness of the neutralizing antibody sotrovimab among high-risk patients with mild-to-moderate SARS-CoV-2 in Qatar. Int J Infect Dis. 2022;124:96–103. https://doi.org/10.1016/j.ijid.2022.09.023.
5. Zheng B, Green ACA, Tazare J, Curtis HJ, Fisher L, Nab L, et al. Comparative effectiveness of sotrovimab and molnupiravir for prevention of severe COVID-19 outcomes in patients in the community: observational cohort study with the OpenSAFELY platform. BMJ. 2022;379:e071932. https://doi.org/10.1136/bmj-2022-071932.
